# Association of pulse pressure with incidence of dementia independent of established risk factors

**DOI:** 10.64898/2026.05.20.26353580

**Authors:** Yijin Fang, Richard N Henson, Richard AI Bethlehem, David J Whiteside, Haoran Zhang, Maria-Eleni Dounavi, Emmanuel A Stamatakis, James B Rowe, Kamen A Tsvetanov

**Author notes:** **Correspondence to:** Yijin Fang, Department of Clinical Neurosciences, University of Cambridge, Cambridge CB2 0QQ, United Kingdom.

## Abstract

**Objectives:** To investigate the association between pulse pressure and dementia incidence, independent of other blood pressure measurements and established risk factors, and to assess whether this association differs across dementia subtypes.

**Design:** Prospective population-based study.

**Setting:** UK Biobank.

**Participants:** Of the 502,211 participants in the UK Biobank, 470,986 completed at least one blood pressure measurement and were included in the analysis. These participants were recruited between March 2006 and July 2010 and were followed for up to four assessments through to February 2024.

**Main outcome measures:** Incidence of dementia, identified through linked health records using ICD-9 and ICD-10 diagnosis codes, self-reported diagnoses or records of dementia-specific medication use.

The association between pulse pressure and risk of dementia was investigated using Cox proportional hazard models. Models were adjusted for age, sex, education, hearing problems, lipid levels, depression, traumatic brain injury, physical activity, diabetes, smoking, hypertension, body mass index, alcohol consumption and mean arterial pressure. Dementia subtype-specific associations were examined using competing risk models, with cause-specific Cox analyses included as supplementary sensitivity analyses.

**Results:** During a median follow-up of 13 years, 9,028 persons developed dementia (Alzheimer’s disease: 3,011; Vascular dementia: 1,270; Dementia with Lewy bodies: 234; Frontotemporal dementia: 191; Other/Mixed dementia: 4,322). Each 10mm Hg increase in pulse pressure was associated with a 5.4% higher risk of dementia (95% confidence interval on hazard ratio: 1.036 to 1.071), even after adjustment for age, mean arterial pressure and other established dementia risk factors. The effects were disproportionately stronger for Alzheimer’s disease and vascular dementia, with no clear evidence for increased risk for dementia with Lewy bodies or frontotemporal dementia. Results were robust across sensitivity analyses including alternative blood pressure metrics, complete-case models, and alternative dementia classifications.

**Conclusions:** Pulse pressure is independently associated with incidence of dementia beyond conventional blood pressure measures.

## Introduction

Dementia affects over 57 million people globally and is a leading cause of disability and mortality, with numbers expected to rise due to population ageing (WHO, 2025). While ageing remains the primary risk factor, growing evidence indicates that a substantial portion of dementia risk is attributable to modifiable factors, particularly related to vascular health (Livingston et al., 2024). Identifying vascular markers that improve early risk stratification is therefore a key public health priority. Hypertension has been consistently identified as a modifiable risk factor for cognitive decline and dementia, supporting blood pressure as an important target for prevention (Kennelly et al., 2009a; Kennelly et al., 2009b). While systolic blood pressure targets are key in cardiovascular disease prevention guidelines, they have not reduced dementia risk or preserved cognition (Levi Marpillat et al., 2013; McGuinness et al., 2009; Rapp et al., 2020). This suggests that there may be another mechanism through which blood pressure affects dementia.

Physiologically, blood pressure comprises a steady-state component (mean arterial pressure) and a pulsatile component (pulse pressure, the difference between systolic and diastolic pressures). Pulse pressure, reflecting arterial stiffness and vascular ageing, may independently contribute to “chronic” brain injury, cognitive decline and dementia (de Montgolfier et al., 2019; King et al., 2025; King et al., 2023; Laosiripisan et al., 2017; Levin et al., 2020; Mohammadi et al., 2025; Nation et al., 2015; Stone et al., 2015; Thorin-Trescases et al., 2018; Wartolowska & Webb, 2022; Zang et al., 2021).However, current evidence derives from relatively small, imaging-based cohort studies, and it remains unclear whether pulse pressure affects dementia risk in large-scale population-based samples, independent of mean arterial pressure and other vascular risk factors. Furthermore, few studies have systematically examined whether pulse pressure shows differential associations across dementia subtypes.

In this prospective cohort study, we examined the association between pulse pressure and dementia incidence, independent of other blood pressure measures and dementia risk factors, in nearly a half a million adults in the UK Biobank (UKB). We then test whether the association between pulse pressure and dementia incidence differs across dementia subtypes.

## Methodology

### Study design

The UKB recruited over 500,000 participants aged 40 to 69 at baseline between March 2006 and July 2010. Participants provided consent for follow-up via linkage to their medical records and the completion of repeated assessments. This research was conducted under UK Biobank application number 20904. The dataset included extensive multimodal information, such as physical and health measures, lifestyle factors, genetic data, medications, and linked electronic health records — including hospital admissions and dementia diagnoses. Data collection has been conducted across up to four assessment waves: baseline (2006–2010), the first repeat assessment (2012–2013), the imaging sub-study (from 2014 onwards), and a second repeat assessment (from 2019 onwards).

As shown in Figure 1, exclusion criteria included participants without valid baseline values for blood pressure, age, sex and dementia status. Participants with baseline dementia, or with diagnosis of dementia at follow up but without a valid diagnosis date, were also excluded. Additionally, participants whose loss-to-follow-up date occurred before their observed exit date (dementia diagnosis, death, or administrative censoring) were excluded. Our final analysis included 469,767 participants (i.e., exclusion of 6.5% of the original sample).

**Figure 1.**
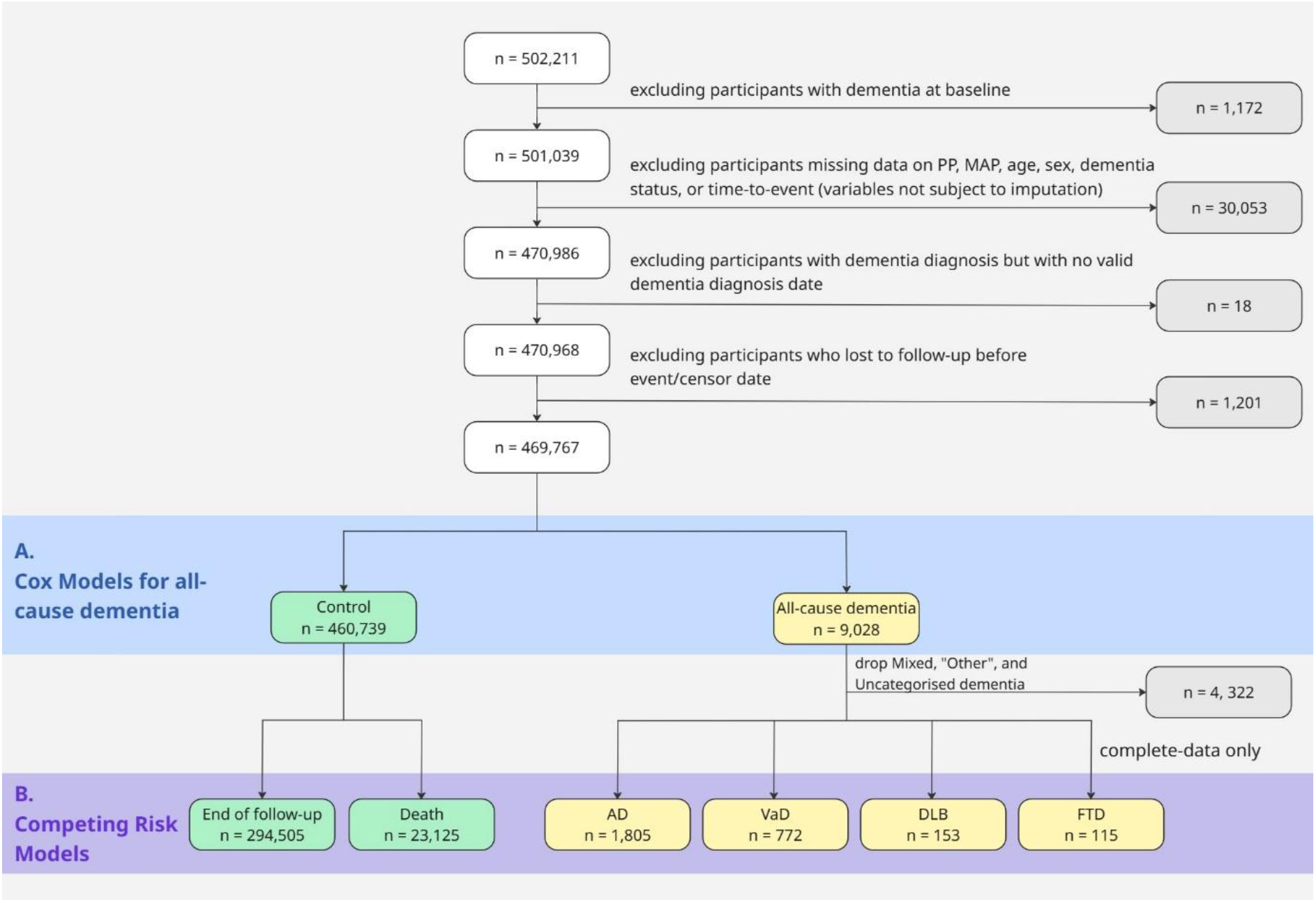
Flowchart of participants inclusion and exclusion across analyses. Participants were drawn from the UK Biobank cohort and sequentially excluded based on baseline dementia status, missing data in key variables not subject to imputation (pulse pressure, mean arterial pressure, age, sex, dementia status, and time-to-event), and missing dementia diagnosis dates. The resulting sample was used for all-cause Cox models (A, frame in light blue). Competing-risk analyses (B, framed in purple) further excluded mixed and uncategorised dementia cases and were conducted in complete-case samples only, excluding participants without complete data on all covariates. Abbreviations: AD, Alzheimer’s disease; VaD, vascular dementia; DLB, dementia with Lewy bodies; FTD, frontotemporal dementia.

### Exposure assessment

In brief, the blood pressure collection procedures followed UKB’s standard protocol. At each assessment instance, systolic and diastolic blood pressure (data-fields 4080 and 4079) were measured twice using an automated device with participants seated at rest, no conversation permitted, and the upper arm unclothed. Between the two readings, participants completed a brief interview and a pulse wave velocity (PWV) measurement. If automated readings were unsuccessful, manual measurements were taken instead (data-fields 93 and 94). Detailed methodology is available on the UKB showcase: https://biobank.ndph.ox.ac.uk/showcase/refer.cgi?id=100225.

From these raw values, three principal variables were computed: pulse pressure, defined as the difference between systolic and diastolic blood pressure, and mean arterial pressure, calculated as diastolic blood pressure plus one-third of pulse pressure (Meaney et al., 2000). These derived measures were used as key vascular predictors in all cross-sectional and longitudinal statistical models. Total blood pressure, the sum of systolic and diastolic blood pressure, was also calculated for sensitivity analysis alternative to mean arterial pressure.

### Ascertainment of outcomes

#### All-cause dementia

Incidence of all-cause dementia was identified using linked hospital inpatient records and self-reported diagnoses, supplemented by medication records indicative of dementia treatment. Detailed information about dementia events in the UK Biobank has been previously published (Anaturk et al., 2023). First, self-report dementia diagnoses (data-field 20002) were identified using UK Biobank codes 1262 (Parkinson’s disease) and 1263 (dementia / Alzheimer’s / cognitive impairment). Second, hospital inpatient records were used, incorporating both ICD-9 (data-field 41271) and ICD-10 (data-field 41270) diagnostic codes. The full list of diagnostic codes used follows the criteria outlined in (Anaturk et al., 2023) and is available in their supplementary materials.

Third, dementia-specific medications (Donepezil, Rivastigmine, Galantamine, Modafinil, and Memantine) were identified via prescription records in data-field 20003. Fourth, additional dementia cases were identified through linked death registry data, including participants with dementia recorded as a primary or secondary cause of death (data-field 40001 and 40002). The same ICD-10 codes were applied as for hospital records.

#### Time-to-dementia incidence

Time-to-incidence variable was calculated to reflect the number of months between each participant’s baseline assessment (instance 0, data-field 53) and the occurrence of a defined event. The event date was defined by: (1) the date of dementia diagnosis, if available; (2) the date of death, if no diagnosis was recorded; or (3) a fixed censoring date at the last date of available linked health records (31 Mar 2023), used for participants with neither a diagnosis nor a recorded death.

When dementia diagnoses were identified from multiple sources (e.g., self-report and hospital records), or multiple records within the same source, the earliest recorded diagnosis date was used to define time to dementia incidence, irrespective of later diagnostic refinement used for subtype classification.

For participants identified solely through death registry records, the date of death was used as the event date in the absence of a recorded diagnostic date.

#### Dementia subtypes

Dementia subtypes were defined using ICD-10 diagnostic codes, given that no dementia cases were identified from ICD-9 records. ICD-10 codes were grouped into the following categories (Lee et al., 2022; Swaddiwudhipong et al., 2023):

1. Alzheimer’s disease: F00, F000, F001, F002, F009, G30, G300, G301, G308, G309
2. Vascular dementia: F01, F010, F011, F012, F013, F018, F019, I673
3. Dementia with Lewy Bodies: F023
4. Frontotemporal dementia: F020, G310

For participants with multiple ICD-10 dementia diagnoses recorded at different time points, the most recent diagnosis record was used to determine dementia subtype.

When multiple dementia diagnoses were recorded on the same most recent date, subtype assignment followed a predefined hierarchical algorithm based on diagnostic specificity and relative prevalence. Specifically, participants were classified as frontotemporal dementia if any frontotemporal dementia diagnosis was present on that date; otherwise, as dementia with Lewy bodies if its diagnosis was present. In the absence of frontotemporal dementia or dementia with Lewy bodies, participants diagnosed with both Alzheimer’s disease and vascular dementia on the same date, with or without an additional diagnosis of other dementia, were classified as having mixed dementia.

Additionally, dementia subtypes through linked death registry data were classified using the same ICD10-codes and hierarchical algorithm described above. Death registry subtype information was used only in the absence of ICD-10 hospital diagnoses; when hospital ICD-10 codes were available, subtype classification was based exclusively on hospital records and death registry codes were not incorporated into the hierarchical assignment.

Participants with specific ICD-10 diagnoses indicating dementia aetiologies other than Alzheimer’s disease, vascular dementia, dementia with Lewy bodies, frontotemporal dementia, or mixed Alzheimer’s disease/ vascular dementia were classified as “Other”, representing rarer clinically defined causes (e.g., other degenerative diseases of nervous system, etc.). Dementia cases identified by self-report or medication records alone, without sufficient diagnostic information to determine subtype, were classified as uncategorised dementia. Due to small sample sizes and/or limited interpretability, mixed, “other”, and uncategorised dementia cases (N=4,372) were excluded from subtype-specific analyses.

#### Covariates

Covariates were selected based on the dementia risk factors identified by Lancet in 2024 that are available in UK Biobank cohort (Livingston et al., 2024). Covariates included age, sex, years of education, hearing problem, low-density lipoprotein (LDL), depression, traumatic brain injury (TBI), physical activity, diabetes, smoking status, hypertension, body matrix index (BMI), and alcohol use. Only the data from instance 0 are used. Reference level for the categorical variables was set as “0” for Cox analyses. The corresponding data-field used for each covariate and the preprocessing steps are summarised in **Error! Reference source not found**.. Detailed information about calculation of years of education in UK Biobank can be found in (Rietveld et al., 2013).

### Statistical Analysis

The participants used for statistical analysis, unless otherwise specified, were filtered to include only those with complete data in blood pressure (systolic and diastolic pressure), age, sex, dementia subtype and time-to-incident, as shown in Figure 1.

#### Descriptive Analyses

Unless otherwise specified, analyses were conducted in participants with complete data on blood pressure measures, age, sex, dementia status, and time-to-incident, as summarised in Figure 1. Differences in demographic and vascular characteristics (age, sex, education, systolic blood pressure, diastolic blood pressure, pulse pressure, mean arterial pressure, total blood pressure and BMI) between dementia cases and controls were assessed using unpaired t-test with Welch correction for continuous variables and Pearson’s chi-squared tests for categorical variable sex. Cohen’s d was calculated as effect size for unpaired t-test using the *effsize* package (Torchiano, 2016). Cramer’s V was calculated as effect size for chi-square test using the formula: 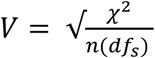 (Tomczak & Tomczak, 2014), where χ is the chi-square statistics, n is the total number of participants for each group, and *df*_*s*_ is the degree of freedom for the smaller number of rows or columns.

#### Cox proportional hazards models

Cox proportional hazards models were used to examine the association between blood pressure variables and dementia incidence. This assumes time-invariant effects of the risk factor. Time-to-event (in months) was defined as the duration from baseline assessment to dementia diagnosis, death, or censoring. All-cause dementia was treated as the event of interest (event = 1), versus an absence of a dementia diagnosis censored at death or the end of follow-up (event = 0).

Pulse pressure was the primary exposure of interest, with mean arterial pressure included to assess independence from steady-state blood pressure components. C-index was used to assess goodness of fit. For clarity, the following abbreviations are used in the model specifications: PP, pulse pressure; MAP, mean arterial pressure; DBP, diastolic blood pressure.

The following models were fitted sequentially:

- *Model 1A: Dementia ∼ PP*
- *Model 1B: Dementia ∼ MAP*
- *Model 1C: Dementia ∼ PP + MAP*
- *Model 1D: Dementia ∼ PP + MAP + Age*
- *Model 1E: Dementia ∼ PP + MAP + Age + Sex + BMI + Years of education + LDL + Smoking status + Alcohol status + Physical activity + Hearing problem + Depression + TBI + Diabetes + Hypertension*

To examine potential moderating effects of selected covariates, Cox proportional hazards models with interaction terms were additionally fitted, in which pulse pressure interacted with one of age, hypertension, sex or diastolic blood pressure, as shown in Models 1F-1I. All models were adjusted for the same covariates as in Model 1E.

- *Model 1F: Dementia ∼ PP + Age + PP:Age + MAP + covariates*
- *Model 1G: Dementia ∼ PP + Hypertension + PP:Hypertension + MAP + covariates*
- *Model 1H: Dementia ∼ PP + Sex + PP:Sex + MAP + covariates*
- *Model 1I: Dementia ∼ PP + DBP + PP:DBP + covariates*

#### Competing risk models

Competing risk models were conducted using R package *cmprsk* (Gray, 2010). To account for the presence of multiple dementia subtype and death as competing events, event status was coded using mutually exclusive categories. Participants censored at the end of follow-up were coded as 0; dementia subtypes were coded as Alzheimer’s disease (1), vascular dementia (2), dementia with Lewy bodies (3), frontotemporal dementia (4); and non-dementia cause of death (5).

Four Fine–Gray subdistribution hazard models were fitted (model 2A – 2D), each treating one dementia subtype as the event of interest and all other dementia subtypes and death as competing events. All models were adjusted for the same covariates as in Model 1E of the Cox proportional hazards analyses. For clarity, the following abbreviations are used in the model specifications: PP, pulse pressure; MAP, mean arterial pressure; AD, Alzheimer’s disease; VaD, vascular dementia; DLB, dementia with Lewy bodies; FTD, frontotemporal dementia.

- *Model 2A (Alzheimer’s Disease as event):* *AD* ∼ *PP* + *MAP* + *covariates, with competing events*: *VaD, DLB, FTD, Death*
- *Model 2B (Vascular dementia as event):* *VaD* ∼ *PP* + *MAP* + *covariates, with competing events*: *AD, DLB, FTD, Death*
- *Model 2C (Dementia with Lewy bodies as event):* *DLB* ∼ *PP* + *MAP* + *covariates, with competing events*: *AD, VaD, FTD, Death*
- *Model 2D (Frontotemporal dementia as event):* *FTD*∼ *PP* + *MAP* + *covariates, with competing events*: *AD, VaD, DLB, Death*

#### Missing data and multiple imputation

While the large sample size allows us to fit the above Cox models to complete cases only, there is always the danger that selecting the subset of participants with complete data leads to biased conclusions (e.g., towards healthier participants). Our default approach was therefore to impute missing values of the covariates mentioned above (dementia status, blood pressure, age and sex were not imputed), including BMI, years of education, LDL, smoking status, alcohol status, physical activity, and hearing problem. Diagnosis-based variables (hypertension, diabetes, depression, and TBI) were coded as binary indicators based on available records (absence of diagnosis coded as 0) and were therefore not included in the imputation procedure. The percentage of total values assumed to be missing at random was 5.3%, while the percentage of participants with at least one missing value was 31.2%. This indicates that a complete-case analysis removing participants with missing value in any covariates would have resulted in a substantial reduction in sample size compared with the current approach, which retains participants with missing covariates through imputation.

The multiple imputation was done by chained equations implemented in the *mice* package in R (Van Buuren & Groothuis-Oudshoorn, 2011), restricted to baseline covariates (instance 0). We planned to exclude participants with fewer than four observed covariates of interest, and to exclude covariates with more than 50% missingness, but no participants nor covariates met these exclusion criteria.

In addition to all other covariates, age and sex were also included as predictors for missing values (but not imputed). Twenty imputed datasets were generated, and Cox analyses were performed within each imputed dataset, with estimates combined using Rubin’s rule. The only exception was for the competing-risk analyses, which were conducted using complete-case data, due to the computation demands of fitting Fine-Gray models for each imputation.

#### Sensitivity analyses

To assess the robustness of observed associations across conventional blood pressure measures, alternative analyses were conducted by replacing mean arterial pressure with systolic blood pressure or total blood pressure. To assess the robustness of the imputation strategy, sensitivity analyses were conducted by refitting the Cox proportional hazard models using complete-case data. Distributions and prevalence of covariates in the imputed datasets were also compared with observed data. Results of the sensitivity and diagnostic analyses are presented in the Supplementary Materials.

To assess the effect of Parkinson disease without dementia, a sensitivity analysis was conducted by refitting the Cox proportional hazard models after removing participants that were identified only with code 1262 from self-report, without any additional dementia codes (1263 from self-report or identified by ICD-9 or 10).

For competing risk models, to assess the impact of handling multiple dementia subtype diagnoses, sensitivity analyses were conducted using alternative subtype definitions. First, analyses were restricted to participants with a single ICD-10 dementia subtype recorded at the most recent diagnosis date, thereby excluding individuals with concurrent multiple subtype diagnoses and removing the need for hierarchical assignment. Second, to evaluate the influence of hierarchical classification, analyses were repeated allowing participants with multiple subtype diagnoses to contribute to each relevant subtype in parallel, by duplicating observations across subtype-specific datasets.

As a supplementary analysis, cause-specific Cox models were conducted within dementia cases only to explore differences in the association between pulse pressure and dementia subtypes. In these models, one subtype was treated as the event of interest (event = 1), while other dementia subtypes were treated as censored observations (event = 0. Non-dementia controls were not included in these analyses. All models were adjusted for the same covariates as in model 1E. Separate models were fitted for each dementia subtype, with the following general model:

- *Model 3A-3D: Dementia subtype (event) ∼ PP + MAP + Age + covariates*

## Results

### Sample characteristics and variable relationships

As summarised in Table 1, the main analyses included 469,767 participants. During a median follow up of 13 years, 9,028 developed dementia after joining the assessments. Compared to controls, individuals with dementia were generally older, had fewer years in education, were slightly more likely to be male, and showed slightly higher systolic blood pressure and BMI on average. Effect size was high for age and pulse pressure, moderate for sex, years of education, mean arterial pressure, systolic blood pressure, and total blood pressure, and small for BMI. The pattern of blood pressure is further illustrated in **Error! Reference source not found**., where dementia cases show a distribution shifted towards higher systolic and lower diastolic blood pressure, consistent with relatively higher pulse pressure.

**Table 1.**
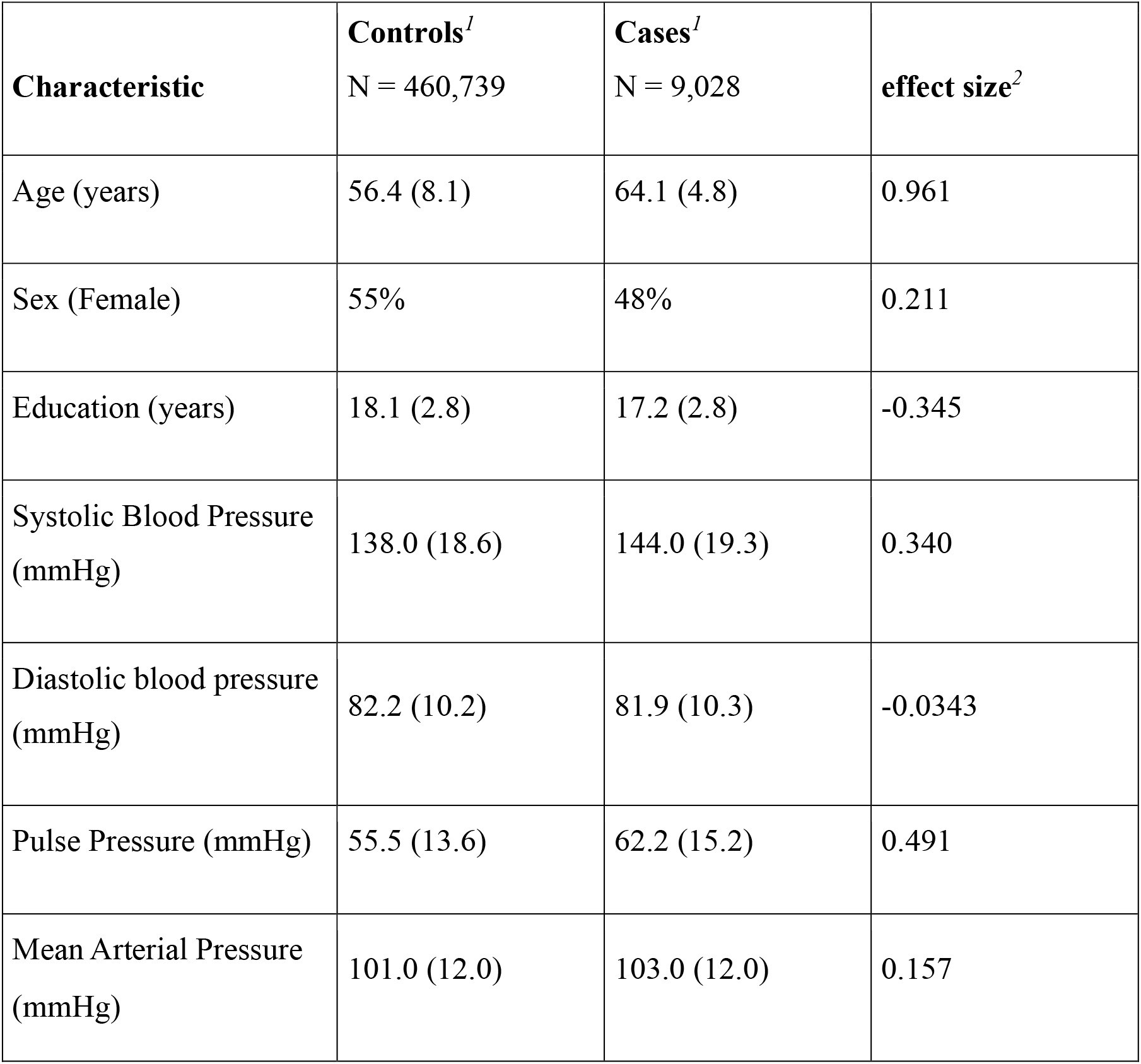
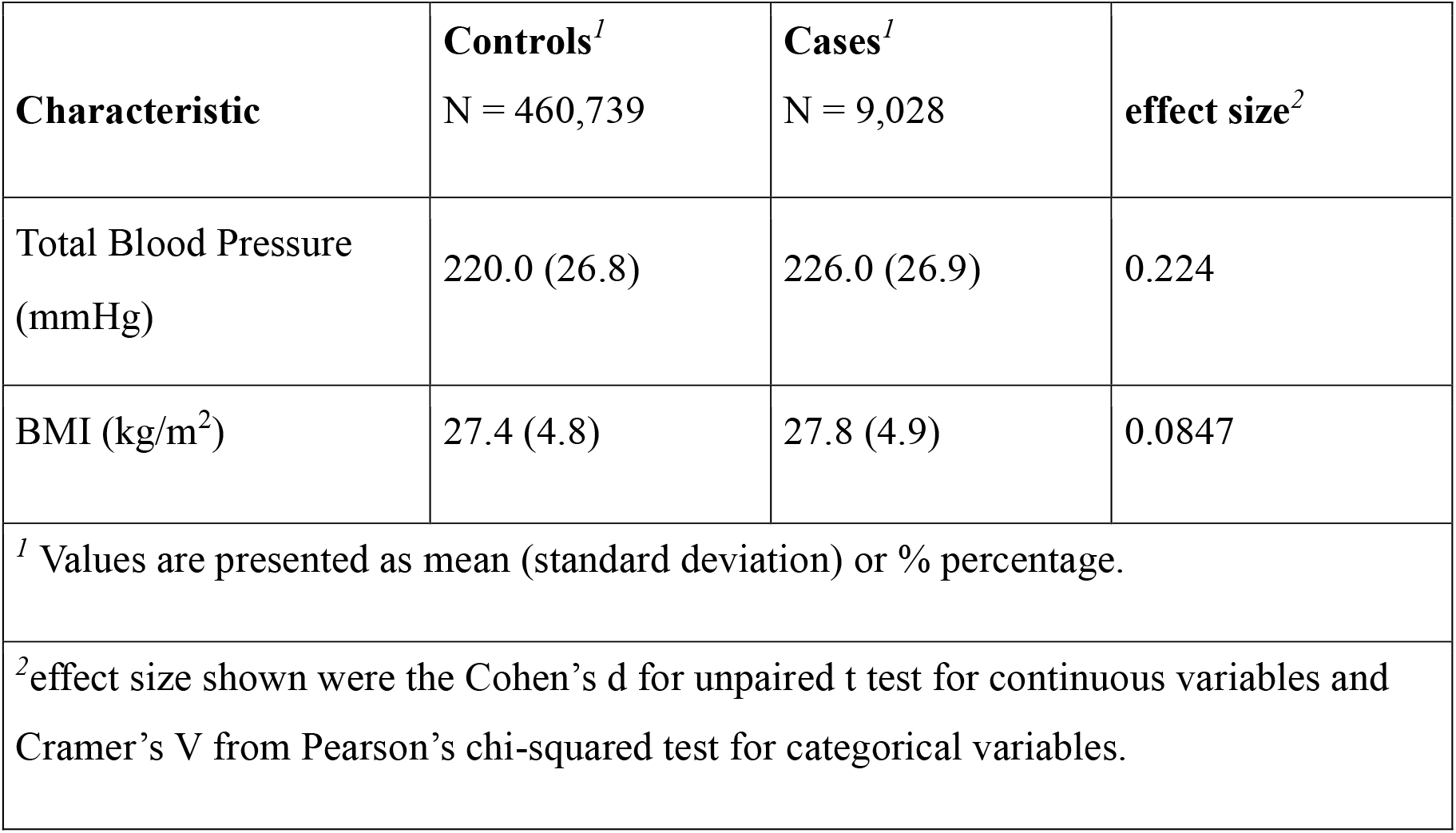
Demographic and Vascular Characteristics of Dementia Cases and Controls.

Figure 2 shows distributions of the main variables (age, pulse pressure, mean arterial pressure, systolic blood pressure, and total blood pressure) and their pairwise correlations. Pulse pressure was moderately correlated with mean arterial pressure (r = 0.560) and total blood pressure (r = 0.671) and strongly correlated with systolic blood pressure (r = 0.847). Pulse pressure also showed a moderate correlation with age (r = 0.417), whereas age was only weakly correlated with mean arterial pressure (r = 0.191). Mean arterial pressure, systolic blood pressure, and total blood pressure exhibited very strong inter-correlations (r > 0.90).

**Figure 2.**
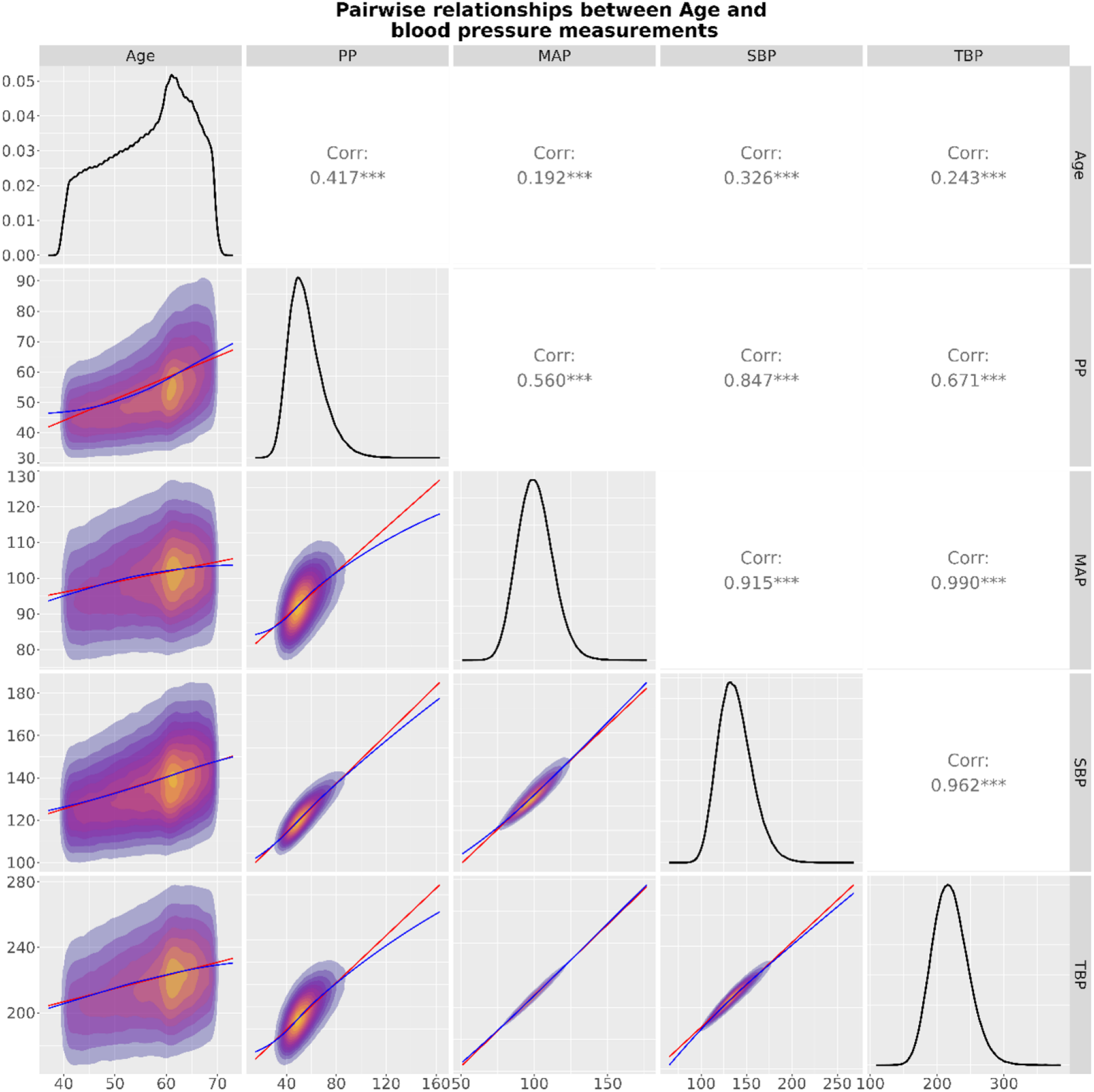
Pairwise relationships between age, pulse pressure, mean arterial pressure, systolic blood pressure, and total blood pressure. Each density plot shows a bivariate distribution of individual participant values, with a fitted linear regression line (red) and a non-parametric smoothing line (blue). The diagonal panels display the histogram of each variable. Pearson’s correlation coefficients are reported in the upper triangle, with asterisks indicating statistical significance (***p < 0.001). Abbreviations: PP, pulse pressure; MAP, mean arterial pressure; SBP, systolic blood pressure; TBP, total blood pressure.

### Cox Proportional Hazard Models

When examined individually, both pulse pressure and mean arterial pressure were positively associated with risk of dementia. Each 10 mmHg increase in pulse pressure was associated with a 31% higher hazard of dementia (HR = 1.031 per 1 mmHg increase, 95% CI 1.030–1.032), while each 10 mmHg increase in MAP corresponded to a 13% higher hazard (HR = 1.013 per 1 mmHg increase, 95% CI 1.012-1.015). See **Error!Reference source not found**. for more details.

When pulse pressure and mean arterial pressure were modelled simultaneously (Model 1C), the association with pulse pressure strengthened (HR = 1.037 per 1 mmHg increase, 95% CI 1.036–1.039), while the association with mean arterial pressure reversed direction (HR = 0.987 per 1 mmHg increase, 95% CI 0.985-0.989).

After further adjustment for age (Model 1D), effect sizes for both pulse pressure and mean arterial pressure were substantially attenuated (HR = 1.006 and 0.995 per 1 mmHg increase, respectively; 95% CI 1.005-1.008 and 0.993-0.997 respectively), while age remained strongly associated with dementia risk (HR = 1.199 per year, 95% CI 1.194-1.205).

With sequential adjustment, model discrimination (C-index) improved, indicating better model fit with the inclusion of additional covariates. Once allowing for all other covariates (Model 1E), each 10 mmHg increase in pulse pressure was associated with a 5.4% higher hazard of dementia, whereas each 10 mmHg increase in mean arterial pressure was associated with a 5.6% lower hazard, as illustrated in Figure 3. The apparent protective association of current alcohol consumption should be interpreted cautiously, as alcohol use was coded categorically and did not reflect quantity of drinking intensity. The observed association is therefore conditional on adjustment for correlated covariates, including previous alcohol use and other health-related factors.

**Figure 3.**
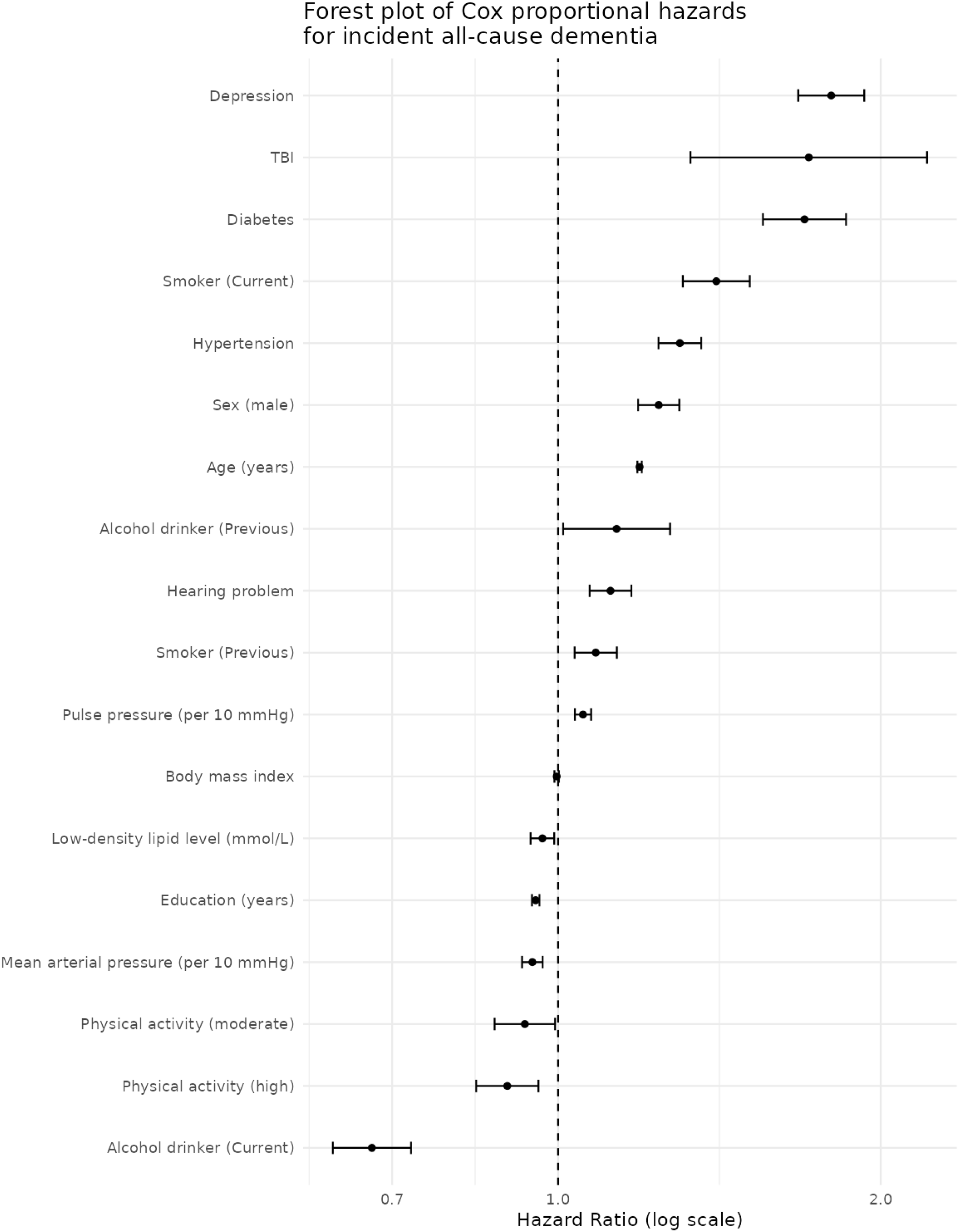
Forest plot of Cox proportional hazards for risk of all-cause dementia from Model 1E. Hazard ratios (HRs) and 95% confidence intervals (CIs). Pulse pressure and mean arterial pressure are expressed per 10 mmHg increase. The vertical dashed line indicates HR = 1. Estimates are plotted on a logarithmic scale. Abbreviations: PP, pulse pressure; MAP, mean arterial pressure.

In models with interactions (Models 1F-I), significant interactions were observed for hypertension status only (**Error! Reference source not found**.). The negative interaction between pulse pressure and hypertension indicated that the association between pulse pressure and dementia risk was stronger among individuals without hypertension and attenuated among those with hypertension. In contrast, there was no evidence that the association between pulse pressure and dementia differed by age, sex, or diastolic blood pressure, indicating that the observed effect of pulse pressure was broadly consistent across these groups.

### Competing-risk Models

Competing-risk models were fitted to examine associations between pulse pressure and the cumulative incidence of dementia subtypes while accounting for competing events, including other dementia subtypes and death. As shown in Table 2, after accounting for competing risks, pulse pressure was moderately associated with the cumulative incidence of Alzheimer’s disease (p = 0.042, with each 10 mmHg increase being associated with a 4% higher hazard), but not with other dementia subtypes.

**Table 2.** Associations between pulse pressure and cumulative incidence of dementia subtype in Competing risk models. The hazard ratio reported for pulse pressure and mean arterial pressure are expressed per 1 mmHg increase. Abbreviations: PP, pulse pressure; MAP, mean arterial pressure; AD, Alzheimer’s disease; VaD, vascular dementia; DLB, dementia with Lewy bodies; FTD, frontotemporal dementia.

| Variables | HR (95% CI) | p-value |
| --- | --- | --- |
| *Model 2A: AD ~ PP + MAP + Age + covariates<sup>1</sup>, with competing events: VaD, DLB, FTD, Death* |  |  |
| PP | 1.004 (1.000-1.008) | 0.042* |
| MAP | 0.996 (0.991-1.001) | 0.120 |
| Age | 1.221 (1.208-1.235) | <0.001 *** |
| *Model 2B: VaD ~ PP + MAP + Age + covariates<sup>1</sup>, with competing events: AD, DLB, FTD, Death* |  |  |
| PP | 1.006 (0.999-1.012) | 0.076 |
| MAP | 0.989 (0.981-0.997) | 0.0091** |
| Age | 1.211 (1.190-1.232) | <0.001 *** |
| *Model 2C: DLB ~ PP + MAP + Age + covariates<sup>1</sup>, with competing events: AD, VaD, FTD, Death* |  |  |
| PP | 0.998 (0.984-1.010) | 0.780 |
| MAP | 1.007 (0.990-1.020) | 0.420 |
| Age | 1.192 (1.155-1.230) | <0.001 *** |
| *Model 2D: FTD ~ PP + MAP + Age + covariates<sup>1</sup>, with competing events: AD, VaD, DLB, Death* |  |  |
| PP | 1.001 (0.987-1.020) | 0.880 |
| MAP | 1.007 (0.989-1.030) | 0.470 |
| Age | 1.067 (1.040-1.100) | <0.001 *** |

<sup>1</sup>*Other covariates adjusted: Sex, BMI, Years of education, LDL, Smoking status, Alcohol status, Physical activity, Hearing problem, depression, TBI, diabetes, and hypertension.*

### Sensitivity Analysis

Replacing mean arterial pressure with systolic blood pressure or total blood pressure yielded effect sizes for pulse pressure that were consistent in direction and larger in magnitude (e.g. HR ranging from 1.0063 to 1.0091 compared with 1.0054 in the primary analysis) in Cox all-cause hazard models (**Error! Reference source not found**.-5). The range of hazard ratios suggest some sensitivity to the choice of correlated blood pressure covariate. In competing risk models, pulse pressure remained associated with Alzheimer’s disease and additionally showed moderate associations with vascular dementia when systolic blood pressure or total blood pressure was used in place of mean arterial pressure (p = 0.013 and 0.038 respectively) (**Error! Reference source not found**.-7).

Exclusion of participants with Parkinson’s disease without dementia did not materially alter results, with changes in hazard ratios for pulse pressure below 0.1% across models (**Error! Reference source not found**.).

Additionally, complete-case Cox analyses produced effect estimates comparable to those obtained from imputed datasets, supporting the robustness of the imputation procedure (**Error! Reference source not found**.), and also suggesting little selection bias in participants with missing values.

Sensitivity analyses using alternative approaches to handling multiple subtype diagnoses yielded broadly consistent results with the primary analysis (**Error! Reference source not found**.-11). Hazard ratios and 95% confidence intervals for pulse pressure were similar across all models. For Alzheimer’s disease, the association with pulse pressure remained borderline significant, with p-values varying slightly across analyses (primary: p = 0.049; single-diagnosis restriction: p = 0.051; multiple-subtype inclusion: p = 0.042). For vascular dementia, the association showed a similar pattern, achieving significance when restricted to single-diagnosis cases (p = 0.049), and showing similar trends in both the primary analysis (p = 0.076) and when allowing multiple subtype classifications (p = 0.072). No significant associations were observed for frontotemporal dementia or dementia with Lewy bodies across any of the analyses.

As a sensitivity analysis to the competing risk models, cause-specific Cox models restricted to dementia cases were fitted, in which each subtype was compared against other dementia subtypes (**Error! Reference source not found**.). In the analysis, pulse pressure was significantly associated with Alzheimer’s disease (HR = 1.005, p < 0.001), but not with vascular dementia, frontotemporal dementia, or dementia with Lewy bodies. This pattern is consistent with the competing risk analyses, in which only Alzheimer’s disease showed a significant association with pulse pressure.

## Discussion

### Statement of principle findings

This study demonstrates a robust association between pulse pressure and incidence of dementia, even after adjustment of mean arterial pressure and established dementia risk factors. Each 10 mmHg increase in pulse pressure was associated with approximately 5% higher hazard of dementia. Stratified analyses suggested that the association between pulse pressure and dementia was stronger among individuals without hypertension.

Across multiple analytic approaches, higher pulse pressure was associated with increased risk of Alzheimer’s disease. Associations with vascular dementia were more consistently observed for mean arterial pressure rather than pulse pressure. Estimates for frontotemporal dementia and dementia with Lewy bodies were imprecise, potentially due to small numbers. No substantial variation in effect estimates was observed across subtypes.

### Strengths and weaknesses of the study

This study has several strengths. It used a large, well-characterised population-based cohort with comprehensive adjustment for conventional blood pressure measures and established modifiable dementia risk factors. Findings were consistent across multiple statistical approaches and sensitivity analysis, including alternative blood pressure specifications and competing risk models, and were robust to multiple imputation of missing covariates.

Several limitations should be acknowledged. The observational design precludes causal inference, and residual confounding from unmeasured or imprecisely measures cannot be excluded. Dementia misclassification is possible, particularly for milder cases and rarer subtypes. Subtype classification may also be influenced by evolving or overlapping pathological processes and by handling of multiple diagnoses. Blood pressure was assessed primarily at baseline and rarely after dementia diagnosis, limiting the ability to capture longitudinal changes and concern for selection bias. UK Biobank cohort is predominantly of White European ancestry, which may limit the generalisability.

### Comparison with other studies

Previous studies have reported associations between pulse pressure and cognitive impairment or dementia risk (King et al., 2025; King et al., 2023; Qiu et al., 2003), although many were limited by smaller samples or did not adjust for other blood pressure measures. The present findings extend this literature by demonstrating an association between pulse pressure and incident dementia after adjustment for mean arterial pressure and established risk factors, suggesting that the pulsatile component of blood pressure contributes information beyond steady-state measures.

The inverse association between mean arterial pressure and dementia, after adjusting for pulse pressure, is also consistent with earlier reports (Verghese et al., 2003) and may reflect the contribution of lower diastolic pressure to reduced cerebral perfusion. Prior studies modelling systolic or diastolic blood pressure categorically or non-linearly have reported subtype-specific associations (Gregson et al., 2019; Lee et al., 2022). Differences in exposure and outcome definitions and modelling strategy likely explain these discrepancies rather than inconsistency in underlying vascular mechanisms.

### Implications and future directions

These findings suggest that the pulsatile component of blood pressure contributes to dementia risk beyond conventional blood pressure measures. Individuals with well-controlled systolic or mean blood pressure may remain at elevated dementia risk if pulse pressure is high, supporting its consideration as a complementary vascular risk marker. The stronger association observed among individuals without hypertension raises the possibility that dementia risk related to elevated pulsatility may be under-recognised in apparently normotensive populations.

The observed differences across dementia subtypes should be interpreted cautiously, and further work is needed to clarify potential differences across dementia types. Future studies should examine non-linear associations and explore underlying mechanisms, including whether elevated pulse pressure contributes to structural brain injury or functional network disruption through microvascular or haemodynamic pathways. Integration with neuroimaging and interventional studies will be important to determine whether targeting pulsatile haemodynamics improves dementia prevention.

## Conclusions

Higher pulse pressure was independently associated with incidence of dementia beyond conventional blood pressure measures and established modifiable risk factors, suggesting it may be an important vascular factor in dementia risk. These findings support further evaluation of the pulsatile component of blood pressure in dementia risk assessment and prevention strategies.

## Data Availability

The data used in this study are available through application to the UK Biobank (https://www.ukbiobank.ac.uk/). Restrictions apply to the availability of these data, which were used under licence for the current study.

## Acknowledgement

Y.F. was supported by the China Scholarship Council (CSC; No. 202408060006). K.A.T. was supported by the Alzheimer’s Society, UK (Grant number 602 and 689). R.N.H. was supported by the UKRI Medical Research Council (SUAG/086 G116768). J.B.R. was supported by the NIHR Cambridge Biomedical Research Centre (NIHR203312: BRC-1215-20014), NIHR funding to the NIHR BioResource (RG94028 & RG85445), Wellcome Trust (220258), Medical Research Council (SUAG/051G101400; SUAG/010 RG91365; MC_UU_00030/14 and MR/T033371/1), the Holt Fellowship and by the Addenbrookes Charitable Trust. M-E.D. was supported by the Alzheimer’s Society, UK (Grant number 612). E.A.S. was supported by the Stephen Erskine Fellow from Queens’ College, Cambridge. We thank UK Biobank participants and staff for their contribution.

## Contributors

Y.F. and K.A.T. conceived and designed the study. Y.F. conducted the analyses and drafted the manuscript. K.A.T. supervised the project and contributed to study design and interpretation. R.N.H. contributed to statistical analyses. J.B.R., R.AI.B., D.J.W., H.Z., M-E.D., R.N.H., and E.A.S. contributed to study interpretation and critically revised the manuscript. All authors approved the final version. Y.F. is the guarantor and accepts full responsibility for the work and the decision to publish.

The corresponding author attests that all listed authors meet authorship criteria and that no others meeting the criteria have been omitted.

## Transparency declaration

The lead author (the manuscript’s guarantor) affirms that this manuscript is an honest, accurate, and transparent account of the study being reported; that no important aspects of the study have been omitted; and that any discrepancies from the study as originally planned have been explained.

## Ethical approval

UK Biobank received ethical approval from the North West Multi-centre Research Ethics Committee (Ref: 21/NW/0157). This research was conducted under UK Biobank application number 20904. Participants provided consent for follow-up via linkage to their medical records and the completion of repeated assessments.

## Data sharing

The data used in this study are available from the UK Biobank upon application by qualified researchers. Statistical code is available from the corresponding author upon reasonable request.

## PPI statement

Patients and the public were not involved in the design, conduct, reporting, or dissemination plans of this research.

