## Supplementary materials for "Association of pulse pressure with incidence of dementia independent of established risk factors"

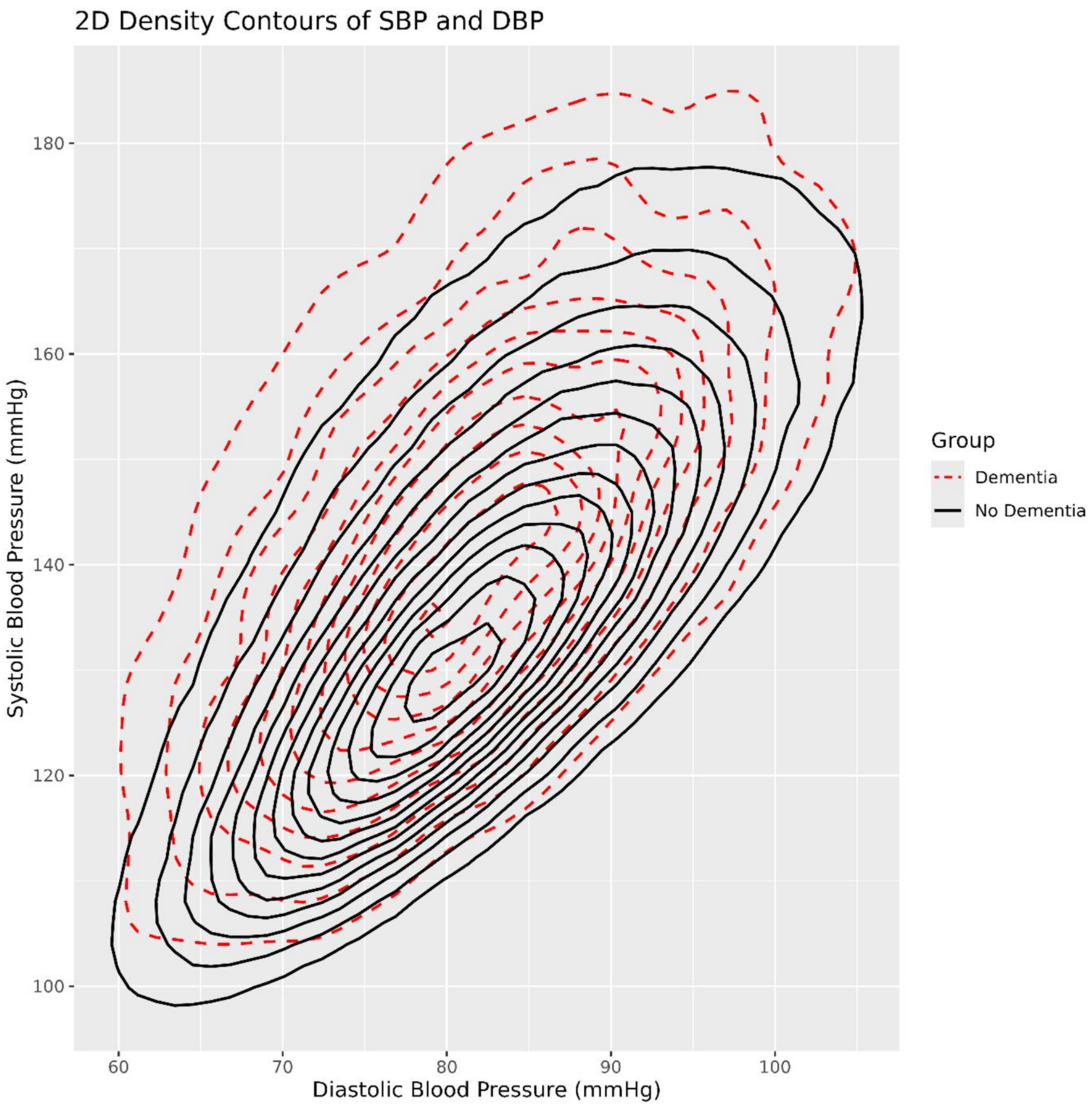


***Supplementary Figure 1. Two-dimensional density contours of systolic and diastolic blood pressure at baseline by dementia status***. Compared with participants without dementia, those who developed dementia showed a distribution shifted towards higher systolic and lower diastolic blood pressure values, consistent with relatively higher pulse pressure. Contours represent kernel density estimates.

***Supplementary Table 1. Preprocessing of confounding factors and covariates. This table*** summarises the confounding factors and covariates included in the Cox regression analyses, their corresponding UK Biobank data-field identifiers, preprocessing procedures, and coding schemes. For diagnostic variables, the specific encoding approaches and ICD-9/ICD-10 codes used for case identification are also provided.

| **Variables** | **Data-field** | **Preprocess** |
|---|---|---|
| Age | 21003 | |
| Sex | 31 | Coding: 0 Female, 1 Male |
| BMI | 21001 | |
| Years of education[1] | 845, 6138 | 1. Identify participant's qualification based on data-field 6138<br>*Coding for data-field 6138: 1 College or University degree; 2 A levels/AS levels or equivalent; 3 O levels/GCSEs or equivalent; 4 CSEs or equivalent; 5 NVQ or HND or HNC or equivalent; 6 Other professional qualifications e.g.: nursing, teaching; -7 None of the above; -3 Prefer not to answer*<br>2. participants without a college degree (coding other than 1): use data field 845, include only positive values from 845<br>3. participants with college degree (coded as 1), assign a value of 21 |
| LDL | 30780 | |
| Smoking Status | 20116 | Coding: 0 never, 1 previous, 2 current, -3 prefer not to answer<br>-3 were converted into NA |
| Alcohol Status | 20117 | Coding: 0 never, 1 previous, 2 current, -3 prefer not to answer<br>-3 were converted into NA |
| Physical Activity | 22032 | IPAQ activity group<br>Coding: 0 for low, 1 for moderate, and 2 for high |
| Hearing problem | 2247 | Coding: 1 for yes, 0 for no, 99 for completely deaf, -1 for do not know, -3 for prefer not to answer<br>99, -1, and -3 were converted into NA |

| | | |
|---|---|---|
| Depression | 20002, 41271, 41270 | 1. Self-report: 1286, 1291, 153<br>2. ICD-9: 3004, 296, 2960-2966, 2968, 311<br>3. ICD-10: F320-F323, F328-F334, F338, F339 |
| TBI | 41271, 41270 | 1. ICD-9: 850-854, 800-804<br>2. ICD-10: S020-S029, S060-S071, S078-S079, S097-S099, T04, T06 |
| Diabetes | 2443, 41271, 41270 | Diabetes: History of Diabetes (Type I or Type II)<br>1. Diabetes was ascertained through self-report (Diabetes diagnosed by doctor)<br>*Coding: 0 No, 1 Yes*<br>2. ICD9: 250, 2500, 2501-2507, 2509<br>3. ICD10: E10, E100-E109, E11, E110-E119, E13, E130-E139, E14, E140-E149 |
| Hypertension | 20002, 41270, 20003 | 1. self report: 1065,1072, 1073<br>2. ICD-10: I10, I11, I12, I13, I15, I674<br>3. Medication: antihypertensive medication (1140888578) |

[1]*detailed information of calculation can be found in previous publications (Rietveld et al., 2013)*

***Supplementary Table 2. Associations between pulse pressure and all-cause dementia risk in Cox proportional hazards models.*** Hazard ratios (HRs), 95% confidence interval (CI), and p-values are shown for stepwise Cox models examining time to risk of all-cause dementia (months) with sequential adjustment for covariates. Results are based on multiply imputed datasets, with estimates combined using Rubin's rules. Reference categories for categorical covariates are provided in the table footnotes. The hazard ratio reported for pulse pressure and mean arterial pressure are expressed per 1 mmHg increase.
*Abbreviations: PP, pulse pressure; MAP, mean arterial pressure.*

| **Variables** | **HR (95% CI)** | **p-value** |
|---|---|---|
| $Model\ 1A: Dementia\ status \sim PP$ (overall model fit[1] = 0.639) | | |
| PP | 1. 031 (1.030–1.032) | <0.001*** |
| $Model\ 1B: Dementia\ status \sim MAP$ (overall model fit[1] = 0.547) | | |
| MAP | 1.013 (1.012-1.015) | <0.001 *** |
| $Model\ 1C: Dementia\ status \sim PP + MAP$ (overall model fit[1] = 0.641) | | |
| PP | 1.037 (1.036-1.039) | <0.001 *** |
| MAP | 0.987 (0.985-0.989) | <0.001 *** |
| $Model\ 1D: Dementia\ status \sim PP + MAP + Age$ (overall model fit[1] = 0.796) | | |
| PP | 1.006 (1.005-1.008) | <0.001 *** |
| MAP | 0.995 (0.993-0.997) | <0.001 *** |
| Age | 1.199 (1.194-1.205) | <0.001 *** |
| $Model\ 1E: Dementia\ status \sim PP + MAP + Age + Sex + BMI + Education\ year + LDL + Smoking\ status + Alcohol\ status + Physical\ activity + Hearing\ problem + depression + TBI + diabetes + hypertension$ (overall model fit[1] = 0.810) | | |
| PP | 1.005 (1.004-1.007) | <0.001 *** |
| MAP | 0.994 (0.992-0.997) | <0.001 *** |
| Age | 1.191 (1.186-1.197) | <0.001 *** |
| Sex (Male) [2] | 1.241 (1.187-1.297) | <0.001 *** |
| BMI | 0.997 (0.992-1.002) | 0.228 |
| Years of education | 0.953 (0.945-0.961) | <0.001 *** |
| LDL | 0.967 (0.942-0.994) | 0.009** |

| | | | |
|---|---|---|---|
| Smoking Status[2] | Previous | 1.084 (1.036-1.134) | <0.001 *** |
| | Current | 1.405 (1.307-1.510) | <0.001 *** |
| Alcohol Status[2] | Previous | 1.134 (1.011-1.272) | 0.032* |
| | Current | 0.670 (0.616-0.729) | <0.001 *** |
| Physical activity[2] | Moderate | 0.931 (0.872-0.993) | 0.031* |
| | High | 0.897 (0.839-0.959) | 0.001** |
| Hearing problem[2] | | 1.119 (1.070-1.170) | <0.001 *** |
| Depression[2] | | 1.798 (1.675-1.930) | <0.001 *** |
| TBI[2] | | 1.713 (1.329-2.290) | <0.001 *** |
| Diabetes[2] | | 1.698 (1.553-1.856) | <0.001 *** |
| Hypertension[2] | | 1.299 (1.241-1.360) | <0.001 *** |

[1]*goodness of model was presented using C-index.*

[2]*Reference categories: female (sex), never smoker, never drinker, low physical activity, no hearing problem, no depression, no traumatic brain injury, no diabetes, and no hypertension.*

***Supplementary Table 3. Interaction effects between pulse pressure and selected covariates on dementia risk in Cox proportional hazards models.*** Hazard ratios (HR) and 95% confidence intervals (CI) are reported for main effects. Interaction effects are presented as both hazard ratios and regression coefficients (β) with corresponding p-values. Models include pulse pressure, mean arterial pressure, and age as core variables (except for the model including diastolic blood pressure where mean arterial pressure is replaced). Each interaction term (pulse pressure × covariate) was examined in a separate model. Negative interaction coefficients indicate attenuation of the association between pulse pressure and dementia with increasing values of the interacting variable. For continuous variables, interaction effects reflect the change in the association per unit increase in the interacting variable.
Abbreviations: PP, pulse pressure; MAP, mean arterial pressure; DBP, diastolic blood pressure.

| **Variables** | **HR (95% CI)** | **β (SE)** | **p-value** |
|---|---|---|---|
| $Model\ 1F: Dementia \sim PP * Age + MAP + covariates$[1] | | | |
| PP | 0.997 (0.977-1.018) | — | 0.797 |
| MAP | 0.995 (0.992-0.997) | — | <0.001 *** |
| Age | 1.183 (1.161-1.205) | — | <0.001 *** |
| PP:Age | 1.000 (1.000-1.000) | $1.246\times10^{-4}$ | 0.446 |
| $Model\ 1F: Dementia \sim PP * hypertension + MAP + Age + covariates$[1] | | | |
| PP | 1.008 (1.005-1.010) | — | <0.001 *** |
| MAP | 0.994 (0.992-0.996) | — | <0.001 *** |
| Age | 1.191 (1.185-1.196) | — | <0.001 *** |
| Hypertension[1] | 1.721 (1.430-2.072) | — | <0.001 *** |
| PP:hypertension | 0.996 (0.993-0.998) | -0.00446 | 0.00214** |
| $Model\ 1F: Dementia \sim PP * Sex + MAP + Age + covariates$[1] | | | |
| PP | 1.007 (1.004-1.009) | — | <0.001 *** |
| MAP | 0.994 (0.992-0.997) | — | <0.001 *** |
| Age | 1.191 (1.186-1.197) | — | <0.001 *** |
| Sex (Male)[1] | 1.439 (1.204-1.720) | — | <0.001 *** |
| PP:Sex | 0.998 (0.995-1.000) | -0.00239 | 0.0927 |
| $Model\ 1F: Dementia \sim PP * DBP + Age + covariates$[1] | | | |
| PP | 1.010 (0.999-1.021) | — | 0.0675 |
| DBP | 1.000 (0.991-1.008) | — | 0.941 |

| | | | |
|---|---|---|---|
| Age | 1.191 (1.186-1.097) | — | <0.001 *** |
| PP:DBP | 1.000 (1.000-1.000) | $-8.180\times10^{-5}$ | 0.226 |

[1]*Other covariates adjusted: Sex, BMI, Years of education, LDL, Smoking status, Alcohol status, Physical activity, Hearing problem, depression, TBI, diabetes, and hypertension.*

***Supplementary Table 4. Associations between pulse pressure and all-cause dementia risk in Cox proportional hazards models (mean arterial pressure replaced with systolic blood pressure).*** Hazard ratios (HRs), 95% confidence interval (CI), and p-values are shown for Cox models examining time to risk of all-cause dementia (months) including all covariates. Results are based on multiply imputed datasets, with estimates combined using Rubin's rules. Reference categories for categorical covariates are provided in the table footnotes. The hazard ratios reported for pulse pressure and systolic blood pressure are expressed per 1 mmHg increase.
Abbreviations: PP, pulse pressure; SBP, systolic blood pressure.

| **Variables** | | **HR (95% CI)** | **p-value** |
|---|---|---|---|
| $Model\ 1E: Dementia\ status \sim PP + SBP + Age + covariates^1$ | | | |
| PP | | 1.009 (1.006-1.012) | <0.001 *** |
| SBP | | 0.994 (0.992-0.997) | <0.001 *** |
| Age | | 1.191 (1.186-1.197) | <0.001 *** |
| Sex (Male) [2] | | 1.241 (1.187-1.297) | <0.001 *** |
| BMI | | 0.997 (0.992-1.002) | 0.228 |
| Years of education | | 0.953 (0.945-0.961) | <0.001 *** |
| LDL | | 0.967 (0.943-0.992) | 0.009** |
| Smoking Status[2] | Previous | 1.084 (1.036-1.134) | <0.001 *** |
| | Current | 1.405 (1.307-1.510) | <0.001 *** |
| Alcohol Status[2] | Previous | 1.134 (1.011-1.272) | 0.032* |
| | Current | 0.670 (0.616-0.729) | <0.001 *** |
| Physical activity[2] | Moderate | 0.931 (0.872-0.993) | 0.031 |
| | High | 0.897 (0.839-0.959) | 0.001** |
| Hearing problem[2] | | 1.119 (1.070-1.170) | <0.001 *** |
| Depression[2] | | 1.798 (1.675-1.930) | <0.001 *** |
| TBI[2] | | 1.713 (1.329-2.209) | <0.001 *** |
| diabetes[2] | | 1.698 (1.553-1.856) | <0.001 *** |
| Hypertension[2] | | 1.299 (1.241-1.360) | <0.001 *** |

[1]*Other covariates adjusted: Sex, BMI, Years of education, LDL, Smoking status, Alcohol status, Physical activity, Hearing problem, depression, TBI, diabetes, and hypertension.*

[2]*Reference categories: female (sex), never smoker, never drinker, low physical activity, no hearing problem, no depression, no traumatic brain injury, no diabetes, and no hypertension.*

***Supplementary Table 5. Associations between pulse pressure and all-cause dementia risk in Cox proportional hazards models (mean arterial pressure replaced with total blood pressure).*** Hazard ratios (HRs), 95% confidence interval (CI), and p-values are shown for Cox models examining time to risk of all-cause dementia (months) including all covariates. Results are based on multiply imputed datasets, with estimates combined using Rubin's rules. Reference categories for categorical covariates are provided in the table footnotes. The hazard ratio reported for pulse pressure and total blood pressure are expressed per 1 mmHg increase. Abbreviations: PP, pulse pressure; TBP, total blood pressure.

| **Variables** | | **HR (95% CI)** | **p-value** |
|---|---|---|---|
| $Model\ 1E: Dementia\ status \sim PP + TBP + Age + covariates$[1] | | | |
| PP | | 1.006 (1.004-1.008) | <0.001 *** |
| TBP | | 0.997 (0.996-0.998) | <0.001 *** |
| Age | | 1.191 (1.186-1.197) | <0.001 *** |
| Sex (Male)[2] | | 1.241 (1.187-1.297) | <0.001 *** |
| BMI | | 0.997 (0.992-1.002) | 0.228 |
| Years of education | | 0.953 (0.945-0.961) | <0.001 *** |
| LDL | | 0.967 (0.943-0.992) | 0.009** |
| Smoking Status[2] | Previous | 1.084 (1.036-1.134) | <0.001 *** |
| | Current | 1.405 (1.307-1.510) | <0.001 *** |
| Alcohol Status[2] | Previous | 1.134 (1.011-1.272) | 0.032* |
| | Current | 0.670 (0.616-0.729) | <0.001 *** |
| Physical activity[2] | Moderate | 0.931 (0.872-0.993) | 0.031 |
| | High | 0.897 (0.839-0.959) | 0.001** |
| Hearing problem[2] | | 1.119 (1.070-1.170) | <0.001 *** |
| Depression[2] | | 1.798 (1.675-1.930) | <0.001 *** |
| TBI[2] | | 1.713 (1.329-2.209) | <0.001 *** |
| diabetes[2] | | 1.698 (1.553-1.856) | <0.001 *** |
| Hypertension[2] | | 1.299 (1.241-1.360) | <0.001 *** |

[1]*Other covariates adjusted: Sex, BMI, Years of education, LDL, Smoking status, Alcohol status, Physical activity, Hearing problem, depression, TBI, diabetes, and hypertension.*

[2]*Reference categories: female (sex), never smoker, never drinker, low physical activity, no hearing problem, no depression, no traumatic brain injury, no diabetes, and no hypertension.*

**Supplementary Table 6. Associations between pulse pressure and cumulative incidence of dementia subtype in competing risk models (mean arterial pressure replaced with systolic blood pressure).** The hazard ratio reported for pulse pressure and systolic blood pressure are expressed per 1 mmHg increase.

Abbreviations: PP, pulse pressure; SBP, systolic blood pressure; AD, Alzheimer's disease; VaD, vascular dementia; DLB, dementia with Lewy bodies; FTD, frontotemporal dementia.

| Variables | HR (95% CI) | p-value |
|---|---|---|
| $Model\ 2A: AD \sim PP + SBP + Age + \ covariates^1,$ $with\ competing\ events: VaD, DLB, FTD, Death$ | | |
| PP | 1.007 (1.000-1.014) | 0.040* |
| SBP | 0.996 (0.991-1.001) | 0.120 |
| Age | 1.221 (1.208-1.235) | <0.001 *** |
| $Model\ 2B: VaD \sim PP + SBP + Age + \ covariates^1,$ $with\ competing\ events: AD, DLB, FTD, Death$ | | |
| PP | 1.013 (1.002-1.023) | 0.013* |
| SBP | 0.989 (0.981-0.997) | 0.00910** |
| Age | 1.211 (1.190-1.232) | <0.001 *** |
| $Model\ 2C: DLB \sim PP + SBP + Age + \ covariates^1,$ $with\ competing\ events: AD, VaD, FTD, Death$ | | |
| PP | 0.993 (0.971-1.020) | 0.560 |
| SBP | 1.007 (0.990-1.020) | 0.420 |
| Age | 1.192 (1.155-1.230) | <0.001 *** |
| $Model\ 2D: FTD \sim PP + SBP + Age + \ covariates^1,$ $with\ competing\ events: AD, VaD, DLB, Death$ | | |
| PP | 0.997 (0.974-1.020) | 0.770 |
| SBP | 1.007 (0.989-1.030) | 0.470 |
| Age | 1.067 (1.040-1.100) | <0.001 *** |

[1]*Other covariates adjusted: Sex, BMI, Years of education, LDL, Smoking status, Alcohol status, Physical activity, Hearing problem, depression, TBI, diabetes, and hypertension.*

***Supplementary Table 7. Associations between pulse pressure and cumulative incidence of dementia subtype in competing risk models (mean arterial pressure replaced with total blood pressure).*** The hazard ratio reported for pulse pressure and total blood pressure are expressed per 1 mmHg increase.
Abbreviations: PP, pulse pressure; TBP, total blood pressure; AD, Alzheimer's disease; VaD, vascular dementia; DLB, dementia with Lewy bodies; FTD, frontotemporal dementia.

| **Variables** | **HR (95% CI)** | **p-value** |
|---|---|---|
| $Model\ 2A: AD \sim PP + TBP + Age + \ covariates^1,$ $with\ competing\ events: VaD, DLB, FTD, Death$ | | |
| PP | 1.005 (1.000-1.010) | 0.037* |
| TBP | 0.998 (0.995-1.001) | 0.120 |
| Age | 1.221 (1.208-1.235) | <0.001 *** |
| $Model\ 2B: VaD \sim PP + TBP + Age + \ covariates^1,$ $with\ competing\ events: AD, DLB, FTD, Death$ | | |
| PP | 1.008 (1.000-1.015) | 0.038* |
| TBP | 0.995 (0.991-0.999) | 0.00910** |
| Age | 1.211 (1.190-1.232) | <0.001 *** |
| $Model\ 2C: DLB \sim PP + TBP + Age + \ covariates^1,$ $with\ competing\ events: AD, VaD, FTD, Death$ | | |
| PP | 0.997 (0.981-1.010) | 0.690 |
| TBP | 1.004 (0.995-1.010) | 0.420 |
| Age | 1.192 (1.155-1.230) | <0.001 *** |
| $Model\ 2D: FTD \sim PP + TBP + Age + \ covariates^1,$ $with\ competing\ events: AD, VaD, DLB, Death$ | | |
| PP | 1.000 (0.984-1.020) | 1.000 |
| TBP | 1.003 (0.994-1.010) | 0.470 |
| Age | 1.067 (1.040-1.100) | <0.001 *** |

[1]*Other covariates adjusted: Sex, BMI, Years of education, LDL, Smoking status, Alcohol status, Physical activity, Hearing problem, depression, TBI, diabetes, and hypertension.*

***Supplementary Table 8. Associations between pulse pressure and all-cause dementia risk in Cox proportional hazards models in participants without Parkinson disease.*** Hazard ratios (HRs), 95% confidence interval (CI), and p-values are shown for stepwise Cox models examining time to risk of all-cause dementia (months) with sequential adjustment for covariates. Results are based on multiply imputed datasets, with estimates combined using Rubin's rules. Reference categories for categorical covariates are provided in the table footnotes. The hazard ratio reported for pulse pressure and mean arterial pressure are expressed per 1 mmHg increase.
Abbreviations: PP, pulse pressure; MAP, mean arterial pressure.

| **Variables** | | **HR (95% CI)** | **p-value** |
|---|---|---|---|
| $Model\ 1A: Dementia\ status \sim PP$ | | | |
| PP | | 1. 031 (1.030–1.033) | <0.001*** |
| $Model\ 1B: Dementia\ status \sim MAP$ | | | |
| MAP | | 1.013 (1.012-1.015) | <0.001 *** |
| $Model\ 1C: Dementia\ status \sim PP + MAP$ | | | |
| PP | | 1.038 (1.036-1.039) | <0.001 *** |
| MAP | | 0.987 (0.985-0.989) | <0.001 *** |
| $Model\ 1D: Dementia\ status \sim PP + MAP + Age$ | | | |
| PP | | 1.006 (1.005-1.008) | <0.001 *** |
| MAP | | 0.995 (0.993-0.997) | <0.001 *** |
| Age | | 1.203 (1.198-1.209) | <0.001 *** |
| $Model\ 1E: Dementia\ status \sim PP + MAP + Age + covariates$[1] | | | |
| PP | | 1.005 (1.004-1.007) | <0.001 *** |
| MAP | | 0.995 (0.992-0.997) | <0.001 *** |
| Age | | 1.195 (1.190-1.201) | <0.001 *** |
| Sex (Male)[2] | | 1.227 (1.174-1.283) | <0.001 *** |
| BMI | | 0.997 (0.992-1.002) | 0.263 |
| Years of education | | 0.951 (0.944-0.959) | <0.001 *** |
| LDL | | 0.965 (0.940-0.991) | 0.008** |
| Smoking Status[2] | Previous | 1.090 (1.042-1.141) | <0.001 *** |
| | Current | 1.432 (1.332-1.539) | <0.001 *** |

| | | | |
|---|---|---|---|
| Alcohol Status[2] | Previous | 1.140 (1.016-1.279) | 0.026* |
| | Current | 0.666 (0.612-0.725) | <0.001 *** |
| Physical activity[2] | Moderate | 0.937 (0.873-1.006) | 0.072 |
| | High | 0.901 (0.838-0.970) | 0.006** |
| Hearing problem[2] | | 1.121 (1.071-1.173) | <0.001 *** |
| Depression[2] | | 1.811 (1.686-1.945) | <0.001 *** |
| TBI[2] | | 1.739 (1.348-2.242) | <0.001 *** |
| Diabetes[2] | | 1.709 (1.562-1.869) | <0.001 *** |
| Hypertension[2] | | 1.309 (1.250-1.371) | <0.001 *** |

[1]*Other covariates adjusted: Sex, BMI, Years of education, LDL, Smoking status, Alcohol status, Physical activity, Hearing problem, depression, TBI, diabetes, and hypertension.*

[2]*Reference categories: female (sex), never smoker, never drinker, low physical activity, no hearing problem, no depression, no traumatic brain injury, no diabetes, and no hypertension.*

***Supplementary Table 9. Associations between pulse pressure and all-cause dementia risk in Cox proportional hazards models in participants with complete data in main predictors and covariates (n total = 322,947; n case = 5,317; n control = 317,630).*** Hazard ratios (HRs), 95% confidence interval (CI), and p-values are shown for stepwise Cox models examining time to risk of all-cause dementia (months) with sequential adjustment for covariates. Results are based on multiply imputed datasets, with estimates combined using Rubin's rules. Reference categories for categorical covariates are provided in the table footnotes. The hazard ratio reported for pulse pressure and mean arterial pressure are expressed per 1 mmHg increase. Abbreviations: PP, pulse pressure; MAP, mean arterial pressure.

| **Variables** | **HR (95% CI)** | **p-value** |
|---|---|---|
| $Model\ 1A: Dementia\ status \sim PP$ | | |
| PP | 1. 032 (1.030–1.034) | <0.001*** |
| $Model\ 1B: Dementia\ status \sim MAP$ | | |
| MAP | 1.014 (1.012-1.016) | <0.001 *** |
| $Model\ 1C: Dementia\ status \sim PP + MAP$ | | |
| PP | 1.038 (1.036-1.040) | <0.001 *** |
| MAP | 0.988 (0.985-0.990) | <0.001 *** |
| $Model\ 1D: Dementia\ status \sim PP + MAP + Age$ | | |
| PP | 1.006 (1.004-1.008) | <0.001 *** |
| MAP | 0.996 (0.993-0.998) | 0.00128** |
| Age | 1.203 (1.196-1.210) | <0.001 *** |
| $Model\ 1E: Dementia\ status \sim PP + MAP + Age + covariates$[1] | | |
| PP | 1.005 (1.003-1.008) | <0.001 *** |
| MAP | 0.994 (0.992-0.997) | <0.001 *** |
| Age | 1.195 (1.188-1.202) | <0.001 *** |
| Sex (Male)[2] | 1.268 (1.197-1.342) | <0.001 *** |
| BMI | 0.996 (0.990-1.003) | 0.228 |
| Years of education | 0.961 (0.951-0.970) | <0.001 *** |
| LDL | 0.974 (0.943-1.005) | 0.103 |
| Smoking Status[2]: Previous | 1.123 (1.060-1.189) | <0.001 *** |
| Smoking Status[2]: Current | 1.430 (1.302-1.571) | <0.001 *** |

| | | | |
|---|---|---|---|
| Alcohol Status[2] | Previous | 1.130 (0.966-1.323) | 0.127 |
| | Current | 0.677 (0.602-0.761) | <0.001 *** |
| Physical activity[2] | Moderate | 0.920 (0.854-0.992) | 0.029* |
| | High | 0.871 (0.807-0.939) | <0.001 *** |
| Hearing problem[2] | | 1.115 (1.054-1.179) | <0.001 *** |
| Depression[2] | | 1.749 (1.592-1.922) | <0.001 *** |
| TBI[2] | | 1.502 (1.055-2.139) | 0.024* |
| Diabetes[2] | | 1.637 (1.452-1.845) | <0.001 *** |
| Hypertension[2] | | 1.311 (1.236-1.391) | <0.001 *** |

[1]*Other covariates adjusted: Sex, BMI, Years of education, LDL, Smoking status, Alcohol status, Physical activity, Hearing problem, depression, TBI, diabetes, and hypertension.*

[2]*Reference categories: female (sex), never smoker, never drinker, low physical activity, no hearing problem, no depression, no traumatic brain injury, no diabetes, and no hypertension.*

**Supplementary Table 10. Associations between pulse pressure and cumulative incidence of dementia subtype in competing risk models (cases restricted to participants with a single ICD-10 dementia subtype recorded only).** The hazard ratio reported for pulse pressure and mean arterial pressure are expressed per 1 mmHg increase. N for controls = 317,630 and n for cases = 2,483.
Abbreviations: PP, pulse pressure; MAP, mean arterial pressure; AD, Alzheimer's disease; VaD, vascular dementia; DLB, dementia with Lewy bodies; FTD, frontotemporal dementia.

| **Variables** | **HR (95% CI)** | **p-value** |
|---|---|---|
| $Model\ 2A: AD \sim PP + MAP + Age + \ covariates^1,$ $with\ competing\ events: VaD, DLB, FTD, Death$ | | |
| PP | 1.004 (1.000-1.009) | 0.051 |
| MAP | 0.996 (0.991-1.002) | 0.160 |
| Age | 1.224 (1.209-1.238) | <0.001 *** |
| $Model\ 2B: VaD \sim PP + MAP + Age + \ covariates^1,$ $with\ competing\ events: AD, DLB, FTD, Death$ | | |
| PP | 1.007 (1.000-1.013) | 0.049* |
| MAP | 0.988 (0.979-0.996) | 0.00410** |
| Age | 1.209 (1.187-1.232) | <0.001 *** |
| $Model\ 2C: DLB \sim PP + MAP + Age + \ covariates^1,$ $with\ competing\ events: AD, VaD, FTD, Death$ | | |
| PP | 0.994 (0.977-1.010) | 0.530 |
| MAP | 1.006 (0.986-1.030) | 0.580 |
| Age | 1.194 (1.147-1.240) | <0.001 *** |
| $Model\ 2D: FTD \sim PP + MAP + Age + \ covariates^1,$ $with\ competing\ events: AD, VaD, DLB, Death$ | | |
| PP | 1.000 (0.986-1.015) | 0.980 |
| MAP | 1.010 (0.991-1.030) | 0.300 |
| Age | 1.067 (1.038-1.096) | <0.001 *** |

[1]*Other covariates adjusted: Sex, BMI, Years of education, LDL, Smoking status, Alcohol status, Physical activity, Hearing problem, depression, TBI, diabetes, and hypertension.*

***Supplementary Table 11. Associations between pulse pressure and cumulative incidence of dementia subtype in competing risk models (participants with multiple subtype diagnoses contributed to each relevant subtype in parallel).*** The hazard ratio reported for pulse pressure and mean arterial pressure are expressed per 1 mmHg increase. N for controls = 317,630 and n for cases = 2,811 (among which 328 participants had multiple dementia subtype diagnosis).
Abbreviations: PP, pulse pressure; MAP, mean arterial pressure; AD, Alzheimer's disease; VaD, vascular dementia; DLB, dementia with Lewy bodies; FTD, frontotemporal dementia.

| **Variables** | **HR (95% CI)** | **p-value** |
|---|---|---|
| $Model\ 2A: AD \sim PP + MAP + Age + covariates^1,$ $with\ competing\ events: VaD, DLB, FTD, Death$ | | |
| PP | 1.004 (1.000-1.009) | 0.042* |
| MAP | 0.996 (0.991-1.002) | 0.170 |
| Age | 1.223 (1.205-1.242) | <0.001 *** |
| $Model\ 2B: VaD \sim PP + MAP + Age + covariates^1,$ $with\ competing\ events: AD, DLB, FTD, Death$ | | |
| PP | 1.006 (0.999-1.012) | 0.072 |
| MAP | 0.989 (0.981-0.997) | 0.00950** |
| Age | 1.212 (1.186-1.237) | <0.001 *** |
| $Model\ 2C: DLB \sim PP + MAP + Age + covariates^1,$ $with\ competing\ events: AD, VaD, FTD, Death$ | | |
| PP | 0.997 (0.982-1.010) | 0.710 |
| MAP | 1.006 (0.989-1.020) | 0.490 |
| Age | 1.194 (1.155-1.240) | <0.001 *** |
| $Model\ 2D: FTD \sim PP + MAP + Age + covariates^1,$ $with\ competing\ events: AD, VaD, DLB, Death$ | | |
| PP | 1.001 (0.987-1.016) | 0.850 |
| MAP | 1.007 (0.988-1.026) | 0.480 |
| Age | 1.068 (1.041-1.097) | <0.001 *** |

[1]*Other covariates adjusted: Sex, BMI, Years of education, LDL, Smoking status, Alcohol status, Physical activity, Hearing problem, depression, TBI, diabetes, and hypertension.*

***Supplementary Table 12. Associations between pulse pressure and risk of dementia subtype in Cause-specific Cox models (cases only).*** Cause-specific Cox models were refitted after removing controls. For each model, the event of interest (coded as 1) was the specified dementia subtype, while participants diagnosed with other dementia subtypes were treated as censored observations (coded as 0). Healthy/non-dementia controls were excluded from these analyses. The hazard ratio reported for pulse pressure and mean arterial pressure are expressed per 1 mmHg increase.
Abbreviations: PP, pulse pressure; MAP, mean arterial pressure; AD, Alzheimer's disease; VaD, vascular dementia; DLB, dementia with Lewy bodies; FTD, frontotemporal dementia.

| **Variables** | **HR (95% CI)** | **p-value** |
|---|---|---|
| $Model\ 3A: AD \sim PP + MAP + Age + covariates$[1] | | |
| PP | 1.005 (1.002-1.008) | <0.001*** |
| MAP | 0.998 (0.994-1.002) | 0.255 |
| Age | 1.006 (0.997-1.015) | 0.174 |
| $Model\ 3B: VaD \sim PP + MAP + Age + covariates$[1] | | |
| PP | 1.004 (0.999-1.008) | 0.125 |
| MAP | 0.996 (0.991-1.002) | 0.203 |
| Age | 1.005 (0.992-1.019) | 0.440 |
| $Model\ 3C: DLB \sim PP + MAP + Age + covariates$[1] | | |
| PP | 1.001 (0.990-1.012) | 0.923 |
| MAP | 1.006 (0.992-1.020) | 0.387 |
| Age | 0.980 (0.951-1.009) | 0.166 |
| $Model\ 3D: FTD \sim PP + MAP + Age + covariates$[1] | | |
| PP | 0.992 (0.979-1.005) | 0.211 |
| MAP | 1.003 (0.989-1.019) | 0.673 |
| Age | 0.899 (0.892-0.934) | <0.001*** |

[1]*Other covariates adjusted: Sex, BMI, Years of education, LDL, Smoking status, Alcohol status, Physical activity, Hearing problem, depression, TBI, diabetes, and hypertension.*
